# End-user preferences and trade-offs for tuberculosis infection testing in a discrete choice experiment: *What do people want?*

**DOI:** 10.64898/2026.05.04.26352357

**Authors:** Amy Hai Yan Chan, Kate Loveys, Matthew Quaife, Alexei Korobitsyn, Nazir Ismail, Assica Hakiman, Ayşenur Kılıç, Yohhei Hamada, Molebogeng X Rangaka

## Abstract

**Introduction:** Tuberculosis infection testing is important but testing uptake is variable. Test-types and attributes may affect engagement, but evidence on end-user values and preferences is lacking. This study aimed to examine end-users preferences and understand trade-offs in test accuracy, operability, cost and convenience.

**Methods:** A cross-sectional discrete choice experiment (DCE) was designed – one for consumers and for health providers. The DCE covered 6 attributes relating test acceptability, accuracy, feasibility and equity. Multinomial logistic regression and willingness-to-accept analyses were used to compare results within and across designs. A minimum sample size of 45 was deemed sufficient to draw statistical inference.

**Results:** A total of 234 people from 59 countries undertook the DCE (48 consumers, 186 providers); 59% female, most from India (26%) and United States (16%). Providers and consumers preferred tests which minimize false positive (β=0.790,p<0.01 consumers; β=0.189,p<0.05 providers) and false negative results (β=0.967,p<0.01; β =0.169,p<0.1). Consumers strongly preferred testing in the community (β=0.395,p=0.07) and in primary care (β=0.427,p=0.02) compared to hospitals. Avoiding in-person follow-up (β=0.151,p=0.02) and not requiring specialist staff or equipment to administer or interpret the test (β=0.144,p=0.01) affected provider preferences. Providers preferred more expensive tests, though overall, test cost was the least important determinant (β=0.133,p=0.05).

Preferences between providers and consumers differed with avoidance of in-person follow-up valued eight times more by providers than consumers. Consumers viewed an increase in likelihood of a false negative result more negatively than an increase in likelihood of a false positive, but the opposite was true among providers.

**Conclusions:** Preferences on TB infection testing was influenced not only by test accuracy but also convenience, portability, and ability to administer in a community / primary care setting. End-users would trade-off key test attributes including test accuracy for convenience. Choice evidence should inform target product profiles for development of novel tests and programmatic implementation.

**Key Messages:**

- What is already known on this topic

- Testing for infection plays a critical role in diagnosing disease and guiding preventive treatment to reduce progression and transmission. However, uptake remains inconsistent, potentially due to test characteristics that do not align with user preferences and expectations.
- What this study adds

- While both providers and consumers value test accuracy, their preferences differ.
- Consumers prioritize accessibility through community or primary care testing, whereas providers emphasize operational simplicity and minimizing in-person follow-up.
- How this study might affect research, practice or policy

- DCE can be implemented across diverse populations to gain broader insights into TB infection testing preferences, emphasizing that test design and delivery should be guided by both provider and consumer perspectives to ensure high uptake and programmatic success.

## Introduction

Tuberculosis is a major cause of health loss (i.e., healthy life years lost to death, illness, or disability), affecting approximately one quarter of the global population and ranked one of the top infectious causes of death^1^. Approximately one quarter to one-third of the global population is estimated to have latent TB infection (TBI) ^2, 3^. Infection may progress to active disease, especially in those with pre-disposing factors such as HIV infection, diabetes, and other immunosuppressive conditions, as well as individuals who are contacts of people with TBI especially children ^1,4^. Testing for infection is important for diagnosis and to guide preventative treatment to curtail progression to active disease and onward disease transmission^5^. However, uptake is variable, affecting uptake implementation.

Current tests for infection include the skin-based purified protein derivative (PPD) tuberculin skin tests (TST) and blood-based interferon gamma release assays (IGRA), which include the WHO-endorsed QuantiFERON-TB Gold (QFT-gold; QFT-gold-in-tube; QFT-plus) and the T-SPOT TB technologies^1^. Despite the importance of TBI testing, there is a large gap in the number of people tested, treated and the numbers estimated to have TBI with treatment coverage averaging about 61%^2^ . This may be due to poor access to TBI testing and / or poor uptake and engagement with TBI testing. These factors can be improved by increasing available options and platforms for TBI testing, improving test performance, and addressing barriers to use. There are many factors that can affect test acceptability and feasibility^3^, including characteristics of the test itself (e.g. invasiveness); test context (e.g. convenience); the individual and health provider (e.g. personal risk perception); and societal factors (e.g. stigma)^4^ ^5^. These factors – or test ‘attributes’ – can influence an individual’s preferences for or against using the test. Understanding the factors that contribute to the acceptability and feasibility of TBI tests can improve utilisation^3^. This is important to inform the development of new tests as well as support the uptake of current tests for infection by identifying the attributes that a desired target product profile of a test for infection of TBI should have.

Identifying how end-user preferences can affect TBI test uptake involves understanding the decision-making processes of the individual. The decision whether to use the test or not is not as straightforward as a simple yes or no; rather the decision often involve a series of ‘trade-offs’ where individuals weigh up their preferences for one attribute over another, selecting the most important attribute for them at the cost of another. For example, in the case of TBI testing, skin tests can be used in community settings though it requires a follow-up for the reading of results; in contrast, IGRA requires blood drawing and laboratory set-up. Trade-off of these attributes can affect the choice of the test and subsequently uptake. A discrete choice experiment (DCE) is a quantitative method that evaluates trade-offs and indirectly elicits stated preferences from participants without directly asking them to state their preferred test options. DCE have been increasingly used in healthcare decision making research and can quantify the strength of user preferences and identify preference heterogeneity across users.

This paper reports the development, testing and findings from a DCE designed to to identify the key attributes and trade-offs that inform user choices and preferences for TBI testing. The objectives of this study were to: conduct a discrete choice experiment (DCE) to explore end-user (consumer and provider) values and preferences for TBI testing; investigate the trade-offs end users are willing to make between test attributes; and determine the order of preference of different test attributes between consumers and health providers.

## Materials and Methods

### Design of DCE tool

#### Attributes and levels

A cross-sectional DCE was designed and conducted online in English. Attributes refer to the features of the test that may influence TBI testing uptake decision and the different types of features that may be available. Levels refer to the values that attributes take in the DCE design. Criteria defined by Hensher et al.^6^ were followed to develop the attributes and levels for the DCE.

Findings from prior literature and qualitative work were used to determine attributes and levels using a standard iterative process^7^. The team first conducted a systematic review identifying all studies that reported end user preferences of TBI tests. The review found that accuracy, convenience, positive patient experience, cost, and resource requirements (such as staff and equipment to administer the test) were important factors influencing test preferences. Second, semi-structured interviews with a mix of 30 consumers (people living with TBI / eligible for TBI testing or underwent testing previously) and providers of TB services (any health provider involved in the TB care pathway) were conducted to identify key test characteristics that would influence user uptake and acceptability of TBI tests. The interviews confirmed the systematic review findings and informed the DCE attributes^5^. Attributes were finalised following input from the World Health Organisation (WHO) Guideline Development Group for “Skin-based tests for *Mycobacterium tuberculosis* antigen-based skin tests”. The group confirmed the key attributes emerging from the qualitative work and literature review, with a focus on policy and clinical importance (Table 1).

**Table 1:**
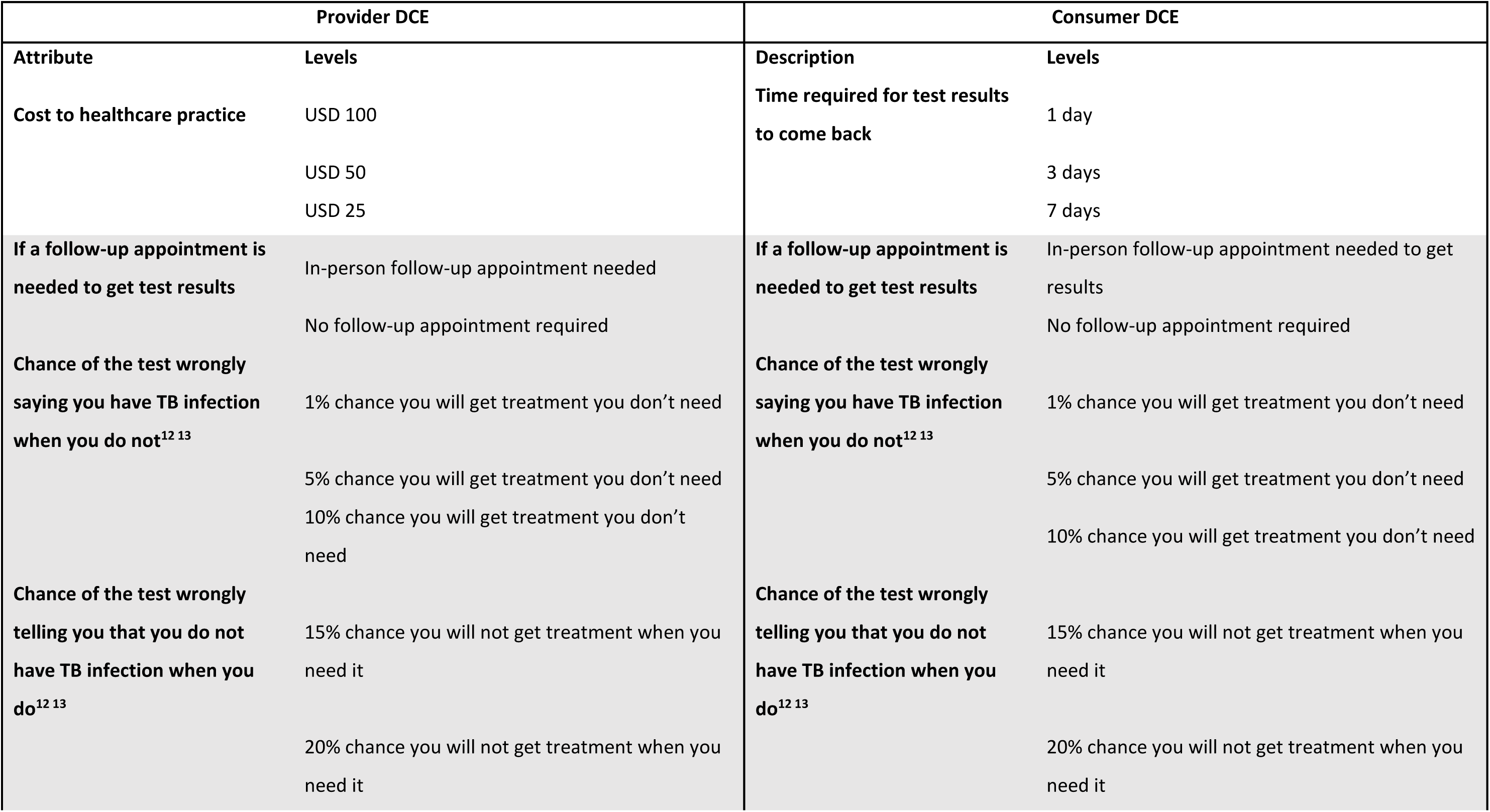
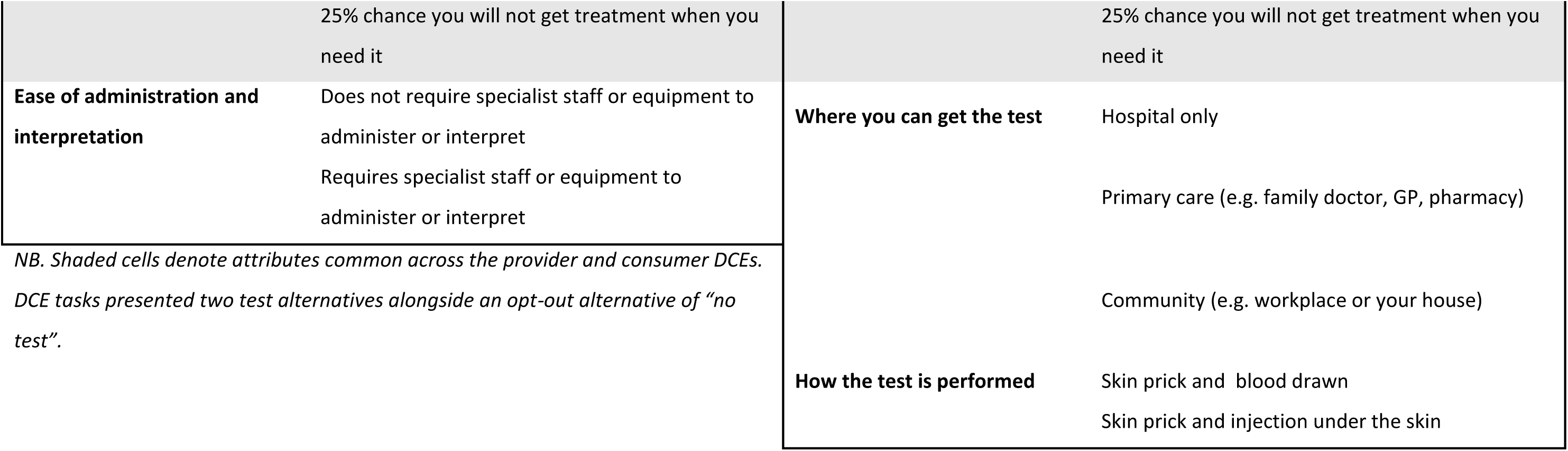
List of attributes and levels included in both provider and consumer DCEs.

Consistent with DCE size in the published literature^8^, we had six attributes with two to six levels for each. The attributes included factors that map to acceptability (waiting time for results, test efficiency, test accuracy (false positives/ false negatives)); feasibility (cost, ease of test administration); and health equity (integration into existing health systems to support equity of access, for example, using existing medical record or reminder systems). Separate DCE choice tasks were created for respondents who were providers of TB services and for respondents who were consumers. Provider and consumer designs differed such that only decision-relevant attributes were shown to each group, for example the provider design included an attribute for staff skill level required, whereas the consumer design included an attribute for location. The full list of attributes included in both DCEs is shown in Table 1.

#### Choice Tasks

Each choice task gave participants a choice between scenarios based on different attribute and level combinations. We constructed a DCE with choice tasks that included two options of hypothetical TB infection test alternatives and an opt-out alternative of choosing neither, which was included to estimate preferences more relevant to real-world choices. The two hypothetical test alternatives are shown in Table 1.

#### Experiment and questionnaire design

A full-factorial design of this size using all the attributes and levels resulted nearly 500,000 possible pairwise choice sets for selection in each DCE. To make tasks manageable to participants, we used NGENE software to construct 10 pairwise choice sets using a D-efficient algorithm. For piloting, we set priors as zero^9^, then used the same software and approach with priors from the pilot to generate a 10-task d-efficient final design. All choice sets were checked for plausibility and dominance. The pilot and final surveys were hosted on an online survey platform (onlinesurveys.ac.uk).

### Participants

Participants were individuals who were either a provider or consumer as defined above. The inclusion criteria were participants ≥18 years, with English at a level sufficient to provide informed consent, and to undertake study procedures including having access to the internet. No prior experience of skin testing or TB was required for inclusion, as the respondents were answering from a hypothetical case scenario perspective. Participants were asked a screener at the start of the DCE: “Are you a provider of TB healthcare?”. Those who answered “yes” received the provider DCE, and those responding with “no” received the consumer DCE. Data were also collected on the gender, age (18-35, 36-55, 56-75, or 75+ years old) and country of the respondent.

### Recruitment

Participants for the DCE were recruited from clinical, research and community networks, including respondents from TB Treatment Action Group, TB Civil Societies, members of the WHO Guideline Development Group for TBST, Global Coalition of TB Activists (GCTA), Stop TB Partnership and the WHO TBI Task Force. Potential participants were provided with an online link, where participants could indicate informed written consent online. Data collection took place between 19th October 2021 and 10^th^ December 2021.

### Data management

After informed consent and self-enrolment on the online survey platform, participants were assigned a unique participant number. All data collected from the participants were labelled using only the participant number to protect participant confidentiality. Participant data were stored in secure password-protected servers at the University of Auckland and LSHTM, and on the onlinesurveys.ac.uk server.

### Ethics statement

An ethics exemption was obtained from our national ethics committee – New Zealand Health and Disability Ethics Committee (HDEC) (21/STH/204). Formal ethics approval was then obtained from the Auckland Health Research Ethics Committee (AH23444).

### Sample size

Diagnostics from the design, specifically the S-error of designs, suggested that statistically significant parameters would be obtained for all attributes in both designs with 45 participants, assuming those in the final sample chose in the same way as the pilot. A minimum sample size of 45 was deemed sufficient to draw statistical inference in both designs, and a recruitment target of approximately 150-200 provider participants and 150-200 consumers was set for the DCE. This sample size target was confirmed after piloting with ten investigators and colleagues to ensure that the DCE gave sufficient information on important test attributes at a sample level^9^.

### Data analysis

Choice data were analysed in a random utility framework using multinomial logit models, and utility weights estimated for the contribution of each attribute level to utility. Dummy coding was used for categorical attributes, and three continuous attributes in each DCE were decided upon at the design stage: the likelihood a test would produce a false positive or negative was modelled as continuous in both provider and non-provider DCEs, while cost was modelled as continuous in the provider DCE, and waiting time in the non-provider DCE.

To explore the trade-offs that participants were willing to make between test attributes, we calculated the marginal rates of substitution between attributes, setting the denominator as the probability of a false positive test result, because this was the most important attribute of the two accuracy attributes common across both DCEs from quantitative analysis of pilot DCE data. The willingness-to-accept results are interpreted as the amount of false-positive accuracy, measured in percentage points of likelihood that a result is a false positive, that participants are willing to forego or bear for the benefit offered by other attributes. To aid interpretation, the cost attribute was set at the central level of USD$50, based on a hypothetical cost, whilst the likelihood of false-positive and false negative results is set at a value of 10% for both based on acceptable diagnostic test criteria^10^.

Where coefficients are significantly greater than zero, an attribute can be interpreted as having a relatively positive impact on participant utility. The magnitude of coefficients can be directly compared within each model. The statistical significance of coefficients (i.e. what is important to people’s choices) can be compared across models, but due to the use of different designs and possible differences in the scale of coefficients across groups, coefficients cannot be directly compared across models. To report the findings of this study, we adapted Discrete Choice Experiment Reporting Checklist (DIRECT)^11^.

## Results

### Characteristics of the study population

In total 234 people responded to the survey, 186 providers and 48 consumers; both sub-populations met the minimum possible sample size needed to draw statistical inference. Table 2 shows the characteristics of these respondents and Figure 1 shows their geographical distribution (see Appendix 1 for full country details). Overall, 59% of respondents were female, the majority (56%) were aged 36-55 years, and the main countries in which respondents were based were the India (26%), United States (16%), South Africa (9%), Pakistan (8%) and Zimbabwe (7%).

**Figure 1:**
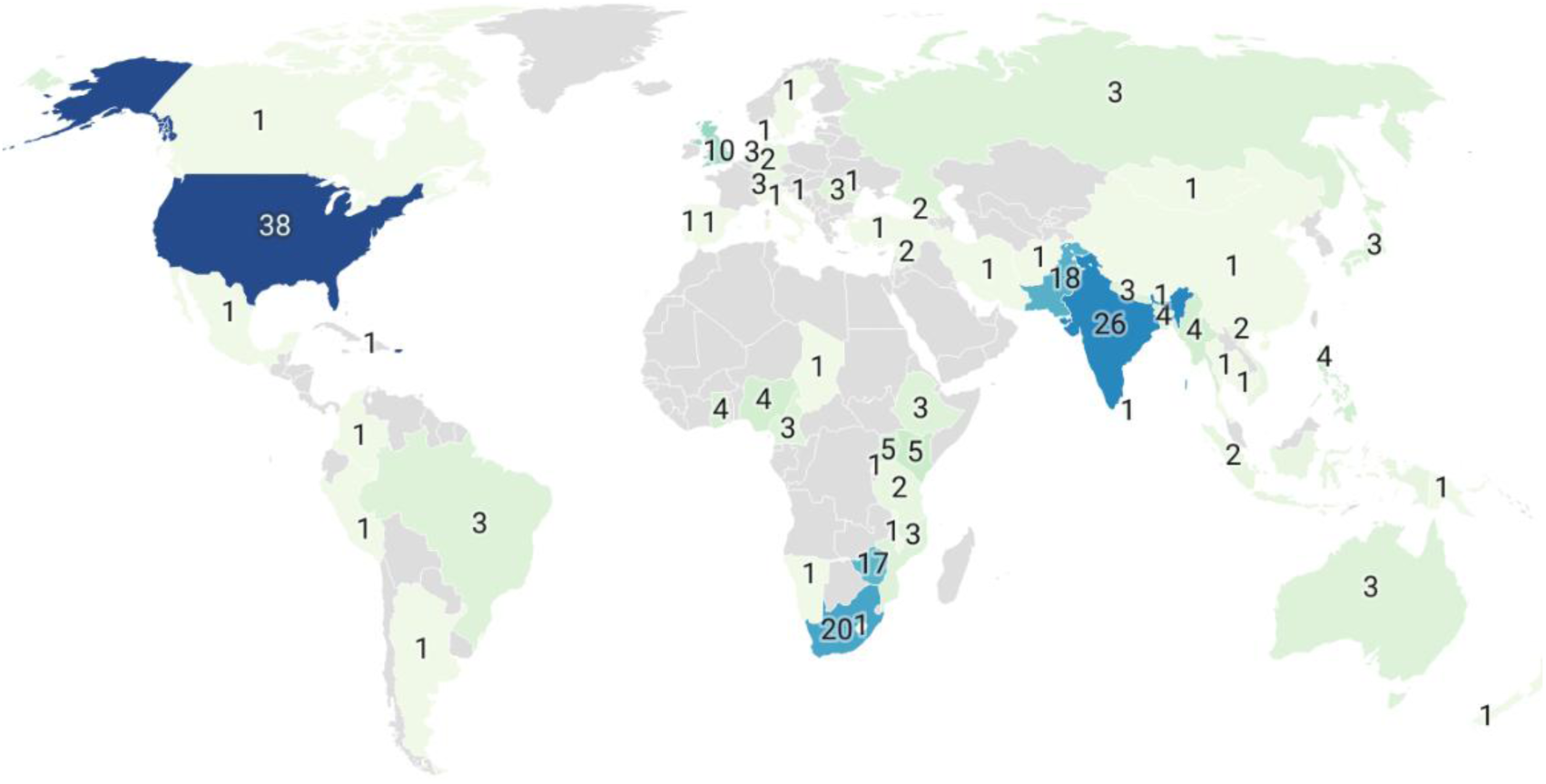
Location of DCE respondents*. *\*Colours represent density of respondents per country, and numbers reflect number of respondents. See Appendix 1 for full details*

**Table 2:** Characteristics of DCE respondents.

|  | **Provider**<br>**(n=186)** | **Consumer (n=48)** |
| --- | --- | --- |
| *Gender* |  |  |
| Female | 112 (60%) | 26 (54%) |
| Male | 73 (39%) | 22 (46%) |
| Other | 1 (1%) |  |
| *Age Category* |  |  |
| 18-35 | 32 (17%) | 13 (27%) |
| 36-55 | 106 (57%) | 24 (50%) |
| 56-75 | 44 (24%) | 11 (23%) |
| 75+ | 4 (2%) | 0 (0%) |

### Choice analysis

In both providers and consumers, the strongest determinant of choice that emerged from the DCE among the evaluated test attributes was test accuracy in terms of false positivity and false negativity rates (Table 3). Preferences for test sensitivity and specificity were not valued differentially among providers (p=0.87) or consumers (p=0.43).

**Table 3:** Multinomial logit choice model results.

|  | (1)<br>Provider |  | (2)<br>Consumer |  |
| --- | --- | --- | --- | --- |
|  | Coefficient | SE | Coefficient | SE |
| No in person follow-up appointment needed | 0.151** | (0.0643) | 0.0829 | (0.175) |
| Chance of the test wrongly saying there is TB infection when there is not (false positive) | -0.189** | (0.0819) | -0.790*** | (0.181) |
| Chance of the test wrongly saying there is no TB infection when there is (false negative) | -0.169* | (0.0960) | -0.967*** | (0.184) |
| Waiting time in days for test result | (not presented as an attribute) |  | -0.0953* | (0.0523) |
| *Place of the test (Reference: hospital)* |  |  |  |  |
| Primary care (e.g. family doctor, GP, pharmacy) | (not presented as an attribute) |  | 0.427** | (0.179) |
| Community (e.g. workplace or your house) |  |  | 0.395* | (0.221) |
| *How the test is performed (Reference: skin prick and blood drawn)* |  |  |  |  |
| Skin prick and injection under the skin | (not presented as an attribute) |  | -0.0599 | (0.262) |
| Cost (USD\$50) | 0.133* | (0.068) | (not presented as an attribute) | |
| Requires specialist staff or equipment to administer or interpret | -0.144** | (0.0562) | (not presented as an attribute) |  |
| Opt-out (no test) | -0.419* | (0.234) | -2.548*** | (0.612) |
| Number of respondents | 186 |  | 48 |  |
| Observations | 5,583 |  | 1,467 |  |
| Bayes Information Criteria | 4096.6 |  | 1015.7 |  |

\*\*\* p<0.01, \*\* p<0.05, \* p<0.1

Among providers, after test accuracy the next most influential attribute was not requiring an in-person follow-up appointment (β=0.151, p=0.02), followed by not requiring specialist staff or equipment to administer or interpret the test (β=0.144, p=0.01). There was weak evidence that providers preferred more expensive tests, though test cost was the least important determinant of choice (β=0.133, p=0.05).

Among consumers, after test accuracy, the next most influential attribute was being able to obtain the test from a primary care setting (e.g. family doctor, pharmacy), which was significantly preferred to obtaining a test from a hospital (β=0.427, p=0.02), whilst community testing (e.g. workplace or home) was weakly preferred to a hospital testing (β=0.395, p=0.07). Consumers displayed no strong preferences for in-person follow up or whether a test required blood to be drawn but had strong and significant preferences for any kind of test compared to no test, indicated by the large and significant negative coefficient for the test opt-out alternative (β=2.55, p<0.01). Cost was not an attribute in the consumer DCE. In both models, almost all coefficients were of the expected sign suggesting good internal validity of results.

### Willingness-to-accept analysis

Figure 2 shows participant willingness to accept a change in false positivity rate for different test and test-related attributes; these coefficient ratios were similar across provider and consumer samples. Willingness to accept a loss of accuracy suggests the alternate attribute is favourable and is indicated by bars in the positive region (above zero), while bars in the negative region below zero indicate that a gain in test accuracy would be needed to compensate for an attribute which is less favourable (i.e. positive values are ‘good’ or preferred outcomes). For example in Panel A, a 7 day wait with the lower negative value is worse than a 3 day wait, which is worse than no wait.

**Figure 2:**
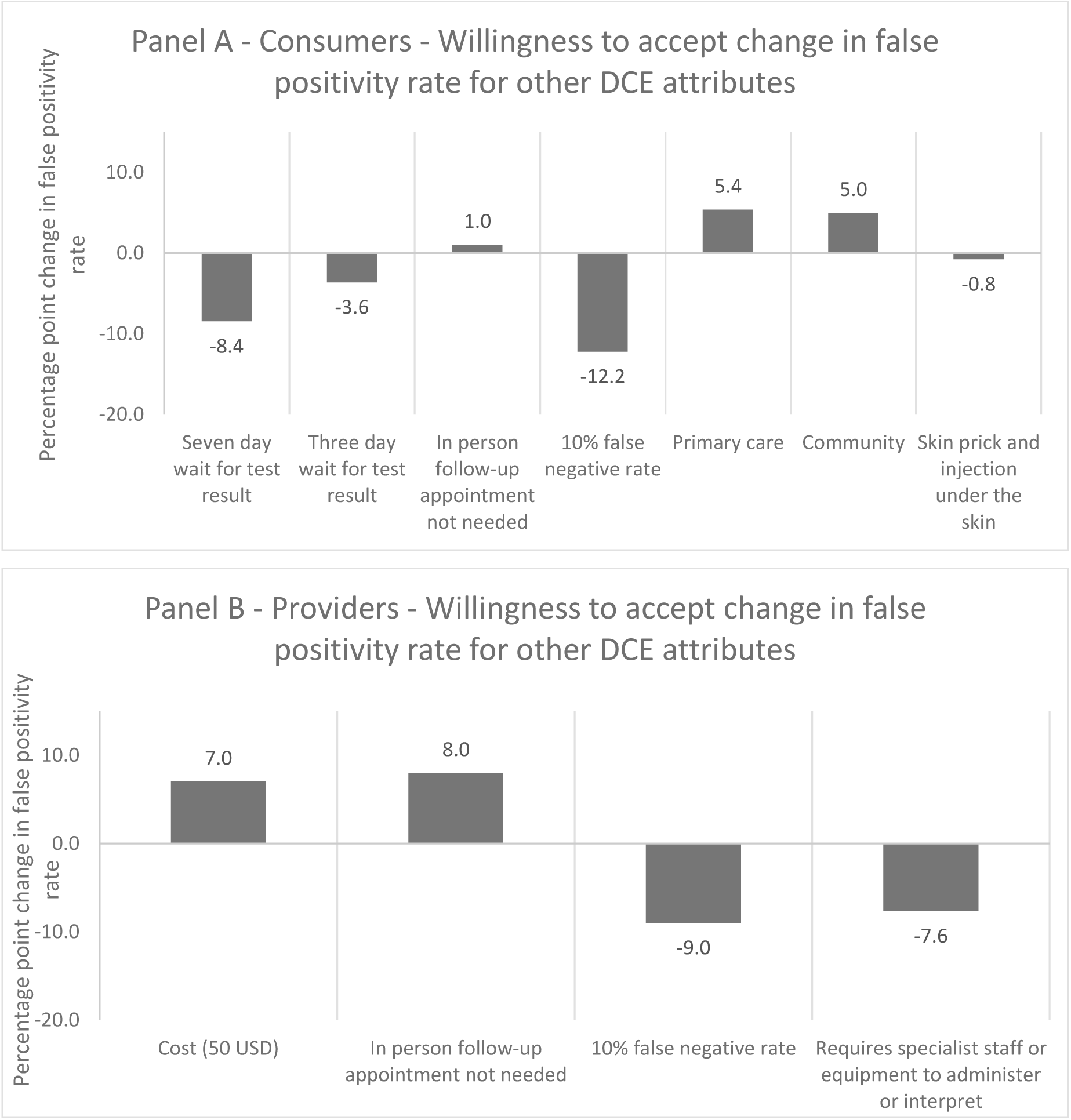
Willingness to accept change in false positivity rate for DCE attributes. Panel A shows results for consumers, and panel B results for providers. A positive value indicates an attribute that is worded ‘positively’ to be a desirable attribute where respondents are willing to accept a lower likelihood of a false-positive result for that attribute. A negative value indicates a ‘negative’ or undesirable attribute that respondents are willing to accept a greater likelihood of a false-positive result for that attribute. Panel A, example interpretation: Consumers would be willing to accept an 8.4 percentage point greater false positivity rate to **avoid** a seven-day wait for test results (an option that is ‘undesirable’ compared to a three-day wait). Panel B, example interpretation: Providers would be willing to forego 8 percentage points of false positivity rate for a test which did not require an in-person follow-up appointment (an option that is ‘desirable’ compared to having to attend an in-person follow-up appointment).

Consumers would be willing to accept an 8.4 percentage point greater false positivity rate to avoid a seven-day wait for test results and a 3.6 percentage point greater rate to avoid a three-day wait, and compared to a one-day wait respectively. Consumers were also willing to forego a 5.4 and 5 percentage point of false positivity rate for a test which did not require a visit to the hospital versus primary care or community setting, respectively (i.e. Primary care and community settings were preferred to hospital settings). The relatively weak consumer preferences for avoiding in-person follow-up and the type of test (skin prick and injection under the skin) are also reflected here (Panel A).

Panel B shows that providers would be willing to forego 8 percentage points of false positivity rate for a test which did not require an in-person follow up appointment, and 7.6 percentage points for a test which does not require specialist staff or equipment to administer or interpret.

There was some evidence of preference heterogeneity between providers and consumers as avoidance of in-person follow-up was valued eight times more by providers than by consumers. Whilst consumers valued a one percentage point increase in the likelihood of a false negative result slightly more negatively than the same increase in likelihood of a false positive; the opposite was true among providers.

## Discussion

This study is the first DCE designed to explore end-user values and preferences with regards to TBI testing. The DCE identified strong and consistent preferences among potential providers and test consumers for tests that minimize false positive and false negative results. Test accuracy was identified as the most important test characteristics by both groups. Following this, consumers’ strongest preferences were for the location of testing, with testing in primary care strongly preferred to hospital locations. For providers, the next most important attribute was the lack of requiring an in-person follow-up appointment, followed by not needing specialist staff or equipment to interpret or administer the test. Willingness-to-accept analyses indicated that both consumers and providers were willing to bear a substantial reduction in test sensitivity to reduce waiting times for results (8.4 percentage point reduction to avoid a 7-day wait), and to avoid in-person follow-up visits (8 percentage points) or the need for specialist staff (7.6 percentage points).

The findings from this DCE highlight the key test attributes that should be considered when it comes to the design of programmes to detect and manage TBI. The findings demonstrate that consumers and providers significantly value convenience attributes, such as where a test is provided and the skill requirements for administering the test and are willing to accept lower test accuracy in return for these benefits. Whilst there is a large body of evidence on the diagnostic accuracy and safety of TBI testing^12^, there remains minimal research into end-user preferences for TBI testing, and factors that influence acceptability and feasibility of testing. For infectious diseases where timely diagnosis is important to limit morbidity and spread of disease, high engagement with testing is paramount.

Our findings provide the first key insights into understanding what factors affect testing uptake, to inform interventions to promote testing. Importantly, the test attributes that were valued by providers differed to that for consumers, except for test accuracy. This is important when considering which test to offer consumers as what may be most acceptable for providers may not be the same for consumers. Interestingly, for test accuracy, it appeared from the DCE trade-offs that consumers would rather receive a false positive result and be treated unnecessarily, than to receive a false negative and not receive treatment and risk having untreated infection. In contrast, the opposite was seen with providers – providers preferred not providing unnecessary treatment (i.e. false positive), and would rather risk a person not getting treatment (i.e. false negative). These differences suggest that consumers prioritize personal health risk, whereas providers focus on clinical responsibility and resource use. These findings may have implications for future TBI test development as the results highlight the need to explore test profiles with potential users, and factors to consider when communicating the benefits of testing to consumers to support shared decision-making that aligns with clinical safety and consumer preferences.

A strength of this study is the use of a DCE methodology to quantitatively assess respondent preferences for tests. The study achieved reasonable geographic diversity in respondents, and results suggest that tasks were understood well by respondents who were able to trade-off over a number of attributes simultaneously. The design process was also strong, designed with multidisciplinary input from global organisations and experts within the field of TBI to design an initial DCE task for piloting. This DCE study has several limitations. First, DCEs require choices between hypothetical alternatives which may not reflect real-world choices accurately or comprehensively. Although there is evidence that DCEs predict real-world choices reasonably well^14^, there remains the potential for hypothetical bias in these data if respondents chose differently in the online survey, than they would have, given different alternatives in real-life. Hence, the impact of different tests on their actual uptake need to be tested in real-world studies. Nevertheless, that preferences align with *a priori* expectations identified from our systematic review and in-depth interviews is an indication of internal validity.

The study was only able to recruit around 25% of the total intended sample size for consumers. However, the coherence and statistical significance of the consumer parameters suggest that our sample size was sufficient to get indicative preference evidence, though this may explain why the consumer sample do not show statistically significant preferences for less important attributes, for example towards requirement for in-person follow-up or how tests are performed. The provider sample was larger, though at 186 was still 14 respondents below the target sample size. There were also few common attributes across the two DCE designs making it difficult to draw conclusions in terms of whether preferences were similar or different in aggregate between providers and consumers – a topic which warrants further future research. Although there was reasonable geographic diversity in responses, there were insufficient responses to explore preference heterogeneity across different geographical settings. To accurately capture how preferences differ by geography, a country- or region-stratified sample is required. Whilst our sample had adequate representation across high, middle and low-income countries, the sample was not large enough to compare preferences across income settings. Our findings may need to be interpreted in the context of the wider health system – higher-income countries with access to specialist staff and equipment, with infrastructure to facilitate follow-up of patients, may have different preferences than low-middle income settings.

## Conclusions

This is the first DCE to explore end-user preferences for TBI testing. Test accuracy, convenience, cost, patient experience and resource requirements were important considerations. Both providers and consumers valued test accuracy. However, consumers ranked portability, and ability to administer in a community / primary care setting as important whilst tests that do not need an in-person follow-up nor specialist staff or equipment to interpret or administer were preferable for health providers. Choice of TBI testing should be guided by both provider and consumer preferences to ensure successful programmatic implementation to improve utilisation and uptake of TBI testing across different healthcare settings.

## Data Availability

Data are available on reasonable request and if allowed by ethics.

## Contributors

AHYC, KL, MQ, AK, NI, YH, and MXR contributed to conceptualisation. AHYC, KL, MQ, YH, MXR did data collection and analysis. AHYC and KL drafted the manuscript and MQ, AK, NI, AH, AK, YH and MXR substantially revised the manuscript. All authors contributed to reviewing and editing the final draft. AHYC is the guarantor.

## Patient and public involvement

Patients and/or the public **were not** involved in the design, or conduct, or reporting, or dissemination plans of this research.

## Provenance and peer review

Not commissioned; externally peer reviewed.

## Author note

I, AHYC, the lead author (the manuscript’s guarantor), affirm that the manuscript is an honest, accurate, and transparent account of the study being reported, that no important aspects of the study have been omitted, and that any discrepancies from the study as planned (and, if relevant, registered) have been explained.

## Supplemental material

N/A

## Data availability statement

Data are available on reasonable request and if allowed by ethics.

## Ethics statements

Patient consent for publication: **Not applicable.**

## Ethics approval

Formal ethics approval was then obtained from the Auckland Health Research Ethics Committee (AH23444)

## Acknowledgements

This work was funded by the World Health Organisation (WHO). This study was part of the WHO guideline group meeting on “Skin-based tests for TB infection, 4-6 February 2022”.

## Appendix 1: Geographical location of DCE respondents

| **Country** | **Provider**<br>**(n=186)** | **Consumer (n=48)** |
| --- | --- | --- |
| United States | 33 | 5 |
| India | 17 | 9 |
| Pakistan | 17 | 1 |
| South Africa | 14 | 6 |
| Zimbabwe | 13 | 4 |
| United Kingdom | 7 | 3 |
| Uganda | 5 |  |
| Kenya | 4 | 1 |
| Bangladesh | 4 |  |
| Philippines | 4 |  |
| Myanmar | 3 | 1 |
| Nigeria | 3 | 1 |
| Brazil | 3 |  |
| Cameroon | 3 |  |
| Japan | 3 |  |
| Romania | 3 |  |
| Russian Federation | 3 |  |
| Ghana | 2 | 2 |
| Australia | 2 | 1 |
| Ethiopia | 2 | 1 |
| Mozambique | 2 | 1 |
| Netherlands | 2 | 1 |
| Switzerland | 2 | 1 |
| Georgia | 2 |  |
| Germany | 2 |  |
| Indonesia | 2 |  |
| Syria | 2 |  |
| Vietnam | 2 |  |
| Tanzania | 1 | 1 |
| Afghanistan | 1 |  |
| Argentina | 1 |  |
| Bhutan | 1 |  |
| Cambodia | 1 |  |
| Canada | 1 |  |
| Chad | 1 |  |
| China | 1 |  |
| Colombia | 1 |  |
| Croatia | 1 |  |
| Denmark | 1 |  |
| Haiti | 1 |  |
| Iran | 1 |  |
| Lesotho | 1 |  |
| Malawi | 1 |  |
| Mexico | 1 |  |
| Moldova | 1 |  |
| Mongolia | 1 |  |
| New Zealand | 1 |  |
| Papua New Guinea | 1 |  |
| Portugal | 1 |  |
| Spain | 1 |  |
| Sweden | 1 |  |
| Thailand | 1 |  |
| Turkey | 1 |  |
| Nepal |  | 3 |
| Italy |  | 1 |
| Namibia |  | 1 |
| Peru |  | 1 |
| Rwanda |  | 1 |
| Sri Lanka |  | 1 |

## Appendix 2: Discrete Choice Experiment Reporting Checklist (DIRECT)

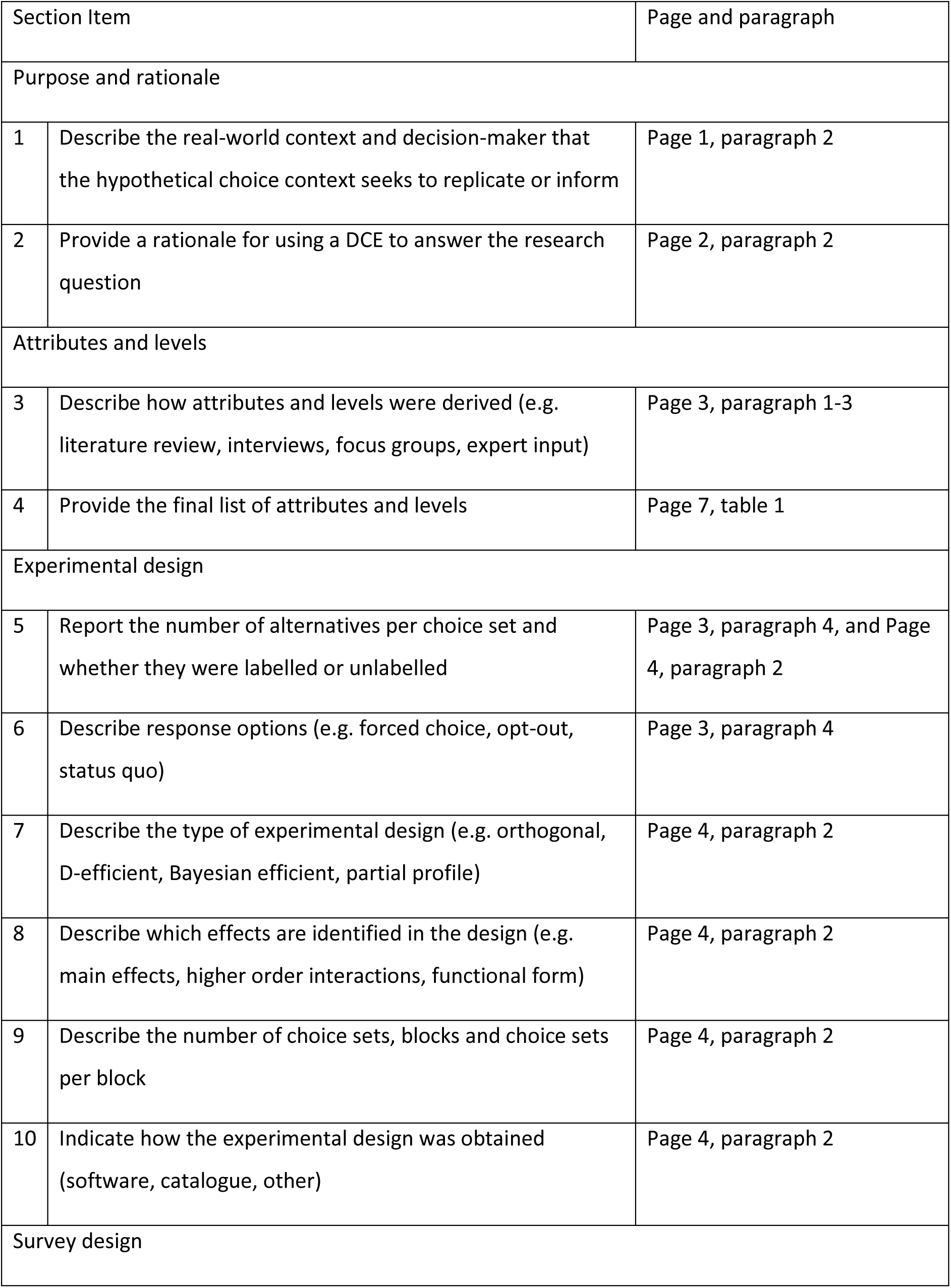
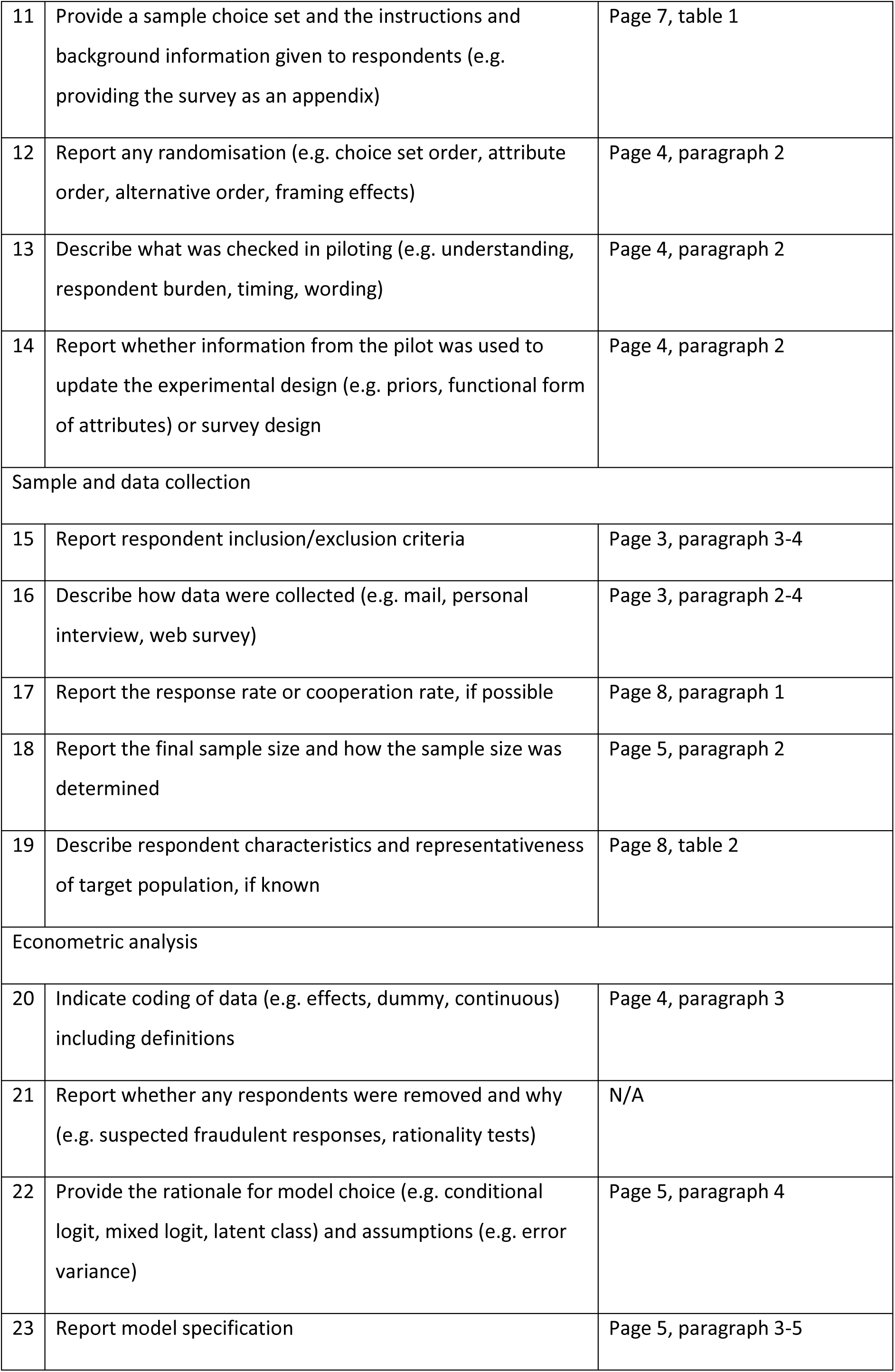
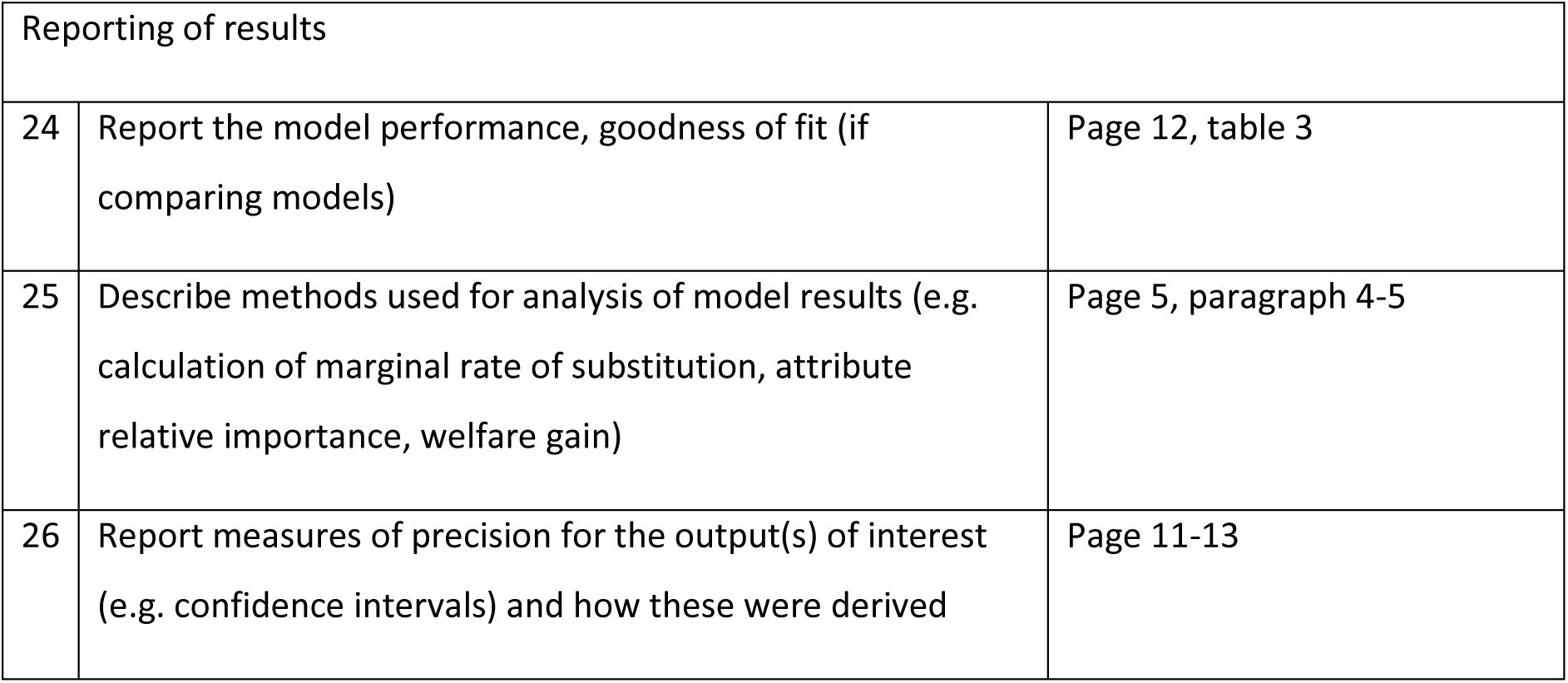

| Section Item |  | Page and paragraph |
| --- | --- | --- |
| Purpose and rationale |  |  |
| 1 | Describe the real-world context and decision-maker that the hypothetical choice context seeks to replicate or inform | Page 1, paragraph 2 |
| 2 | Provide a rationale for using a DCE to answer the research question | Page 2, paragraph 2 |
| Attributes and levels |  |  |
| 3 | Describe how attributes and levels were derived (e.g. literature review, interviews, focus groups, expert input) | Page 3, paragraph 1-3 |
| 4 | Provide the final list of attributes and levels | Page 7, table 1 |
| Experimental design |  |  |
| 5 | Report the number of alternatives per choice set and whether they were labelled or unlabelled | Page 3, paragraph 4, and Page 4, paragraph 2 |
| 6 | Describe response options (e.g. forced choice, opt-out, status quo) | Page 3, paragraph 4 |
| 7 | Describe the type of experimental design (e.g. orthogonal, D-efficient, Bayesian efficient, partial profile) | Page 4, paragraph 2 |
| 8 | Describe which effects are identified in the design (e.g. main effects, higher order interactions, functional form) | Page 4, paragraph 2 |
| 9 | Describe the number of choice sets, blocks and choice sets per block | Page 4, paragraph 2 |
| 10 | Indicate how the experimental design was obtained (software, catalogue, other) | Page 4, paragraph 2 |
| Survey design |  |  |
| 11 | Provide a sample choice set and the instructions and background information given to respondents (e.g. providing the survey as an appendix) | Page 7, table 1 |
| 12 | Report any randomisation (e.g. choice set order, attribute order, alternative order, framing effects) | Page 4, paragraph 2 |
| 13 | Describe what was checked in piloting (e.g. understanding, respondent burden, timing, wording) | Page 4, paragraph 2 |
| 14 | Report whether information from the pilot was used to update the experimental design (e.g. priors, functional form of attributes) or survey design | Page 4, paragraph 2 |
| Sample and data collection |  |  |
| 15 | Report respondent inclusion/exclusion criteria | Page 3, paragraph 3-4 |
| 16 | Describe how data were collected (e.g. mail, personal interview, web survey) | Page 3, paragraph 2-4 |
| 17 | Report the response rate or cooperation rate, if possible | Page 8, paragraph 1 |
| 18 | Report the final sample size and how the sample size was determined | Page 5, paragraph 2 |
| 19 | Describe respondent characteristics and representativeness of target population, if known | Page 8, table 2 |
| Econometric analysis |  |  |
| 20 | Indicate coding of data (e.g. effects, dummy, continuous) including definitions | Page 4, paragraph 3 |
| 21 | Report whether any respondents were removed and why (e.g. suspected fraudulent responses, rationality tests) | N/A |
| 22 | Provide the rationale for model choice (e.g. conditional logit, mixed logit, latent class) and assumptions (e.g. error variance) | Page 5, paragraph 4 |
| 23 | Report model specification | Page 5, paragraph 3-5 |

| Reporting of results |  |  |
| --- | --- | --- |
| 24 | Report the model performance, goodness of fit (if comparing models) | Page 12, table 3 |
| 25 | Describe methods used for analysis of model results (e.g. calculation of marginal rate of substitution, attribute relative importance, welfare gain) | Page 5, paragraph 4-5 |
| 26 | Report measures of precision for the output(s) of interest (e.g. confidence intervals) and how these were derived | Page 11-13 |

